# Silence is golden, by my measures still see: why cheap-but-noisy outcome measures can be more cost effective than gold standards

**DOI:** 10.1101/2022.05.17.22274839

**Authors:** Benjamin Woolf, Hugo Pedder, Henry Rodriguez-Broadbent, Phil Edwards

## Abstract

**Objective:** To assess the cost-effectiveness of using cheap-but-noisy outcome measures, such as a short and simple questionnaire.

**Background:** To detect associations reliably, studies must avoid bias and random error. To reduce random error, we can increase the size of the study and increase the accuracy of the outcome measurement process. However, with fixed resources there is a trade-off between the number of participants a study can enrol and the amount of information that can be collected on each participant during data collection.

**Method:** To consider the effect on measurement error of using outcome scales with varying numbers of categories we define and calculate the Variance from Categorisation that would be expected from using a category midpoint; define the analytic conditions under-which such a measure is cost-effective; use meta-regression to estimate the impact of participant burden, defined as questionnaire length, on response rates; and develop an interactive web-app to allow researchers to explore the cost-effectiveness of using such a measure under plausible assumptions.

**Results:** Compared with no measurement, only having a few categories greatly reduced the Variance from Categorization. For example, scales with five categories reduce the variance by 96% for a uniform distribution. We additionally show that a simple measure will be more cost effective than a gold-standard measure if the relative increase in variance due to using it is less than the relative increase in cost from the gold standard, assuming it does not introduce bias in the measurement. We found an inverse power law relationship between participant burden and response rates such that a doubling the burden on participants reduces the response rate by around one third. Finally, we created an interactive web-app (https://benjiwoolf.shinyapps.io/cheapbutnoisymeasures/) to allow exploration of when using a cheap-but-noisy measure will be more cost-effective using realistic parameter.

**Conclusion:** Cheap-but-noisy questionnaires containing just a few questions can be a cost effect way of maximising power. However, their use requires a judgment on the trade-off between the potential increase in risk information bias and the reduction in the potential of selection bias due to the expected higher response rates.

**Key Messages:**

- A cheap-but-noisy outcome measure, like a short form questionnaire, is a more cost-effective method of maximising power than an error free gold standard when the percentage increase in noise from using the cheap-but-noisy measure is less than the relative difference in the cost of administering the two alternatives.
- We have created an R-shiny app to facilitate the exploration of when this condition is met at https://benjiwoolf.shinyapps.io/cheapbutnoisymeasures/
- Cheap-but-noisy outcome measures are more likely to introduce information bias than a gold standard, but may reduce selection bias because they reduce loss-to-follow-up. Researchers therefore need to form a judgement about the relative increase or decrease in bias before using a cheap-but-noisy measure.
- We would encourage the development and validation of short form questionnaires to enable the use of high quality cheap-but-noisy outcome measures in randomised controlled trials.

## Introduction

Good clinical trials are those that answer ‘important’ questions reliably.(1) An ‘important’ question is one that has not yet been answered reliably, which implies that the size of the effect of the treatment on the disease is likely to be moderate. In order to detect moderate treatment effects in clinical trials and other study designs, systematic errors (bias) and random errors must be avoided.(2) Bias can be avoided by ensuring that treatment allocation is randomised and properly concealed, that outcome assessment is blind to treatment allocation, and that an intention-to-treat analysis is conducted without undue emphasis on subgroups of patients.(3) Random error can be reduced by increasing the size of the study (i.e., reducing sampling error) and by increasing the accuracy of the measurement process (i.e., reducing measurement error).(4–6)

When resources are fixed there is a trade-off between the number of participants a study can enrol and the amount of information that can be collected on each participant during data collection. Since it is only possible to improve the accuracy of a measure by a finite amount, some important questions will need large clinical trials.(1,2) In large studies, collecting outcome data by telephone interview, postal or online questionnaires may be the only financially viable options. For example, if £50,000 was available for outcome data collection and the cost of a detailed assessment of outcome by a trained nurse or doctor was £50 per patient, it would be possible to assess outcomes on 1,000 patients; If the cost of sending a questionnaire and two reminders to each patient or their carer was £5 per patient, it would be possible to assess outcomes on 10,000 patients.

Questionnaire response is associated with participant burden, i.e. amount of effort required by a participant to respond to the questionnaire; For example, the odds of a participant providing outcome data are 60% greater (OR = 1.61, 95% CI: 1.36 to 1.89) using a shorter questionnaire.(7) However, although cheaper, short questionnaires may contain more measurement errorthan the gold standard, and therefore not improve study power despite a larger sample size. They also may not accurately measure the outcome of interest, hence the importance of using scales / measurements that have been properly validated.

In Psychology, unlike Medicine, diagnoses are typically based on measurement instruments rather than clinical judgment. In classical Psychometrics, a common way of constructing instruments is by using questionnaires requiring either yes/no responses or ‘Likert scales’. Likert scales are traditionally questions with 5 (or 3) levels of response (e.g. “strongly agree”, “agree”, “neutral”, “disagree”, “strongly disagree”) which are each given a numerical value in analysis. Although technically an ordered categorical variable, the summation of many Likert questions tends to approximate a normal distribution, and are therefore treated as ordinal or ratio scales.(8,9) Computerised Likert scales sometimes avoid this issue by replacing the categorical question with a continuous scale. If these instruments are valid, then using them as a measure of the continuous underlying liability of the outcome will have greater power than an analysis which dichotomised the trait based on a clinical threshold.(10) For disorders in which a “gold standard” measure, such as a doctor’s diagnosis or biometric measurement is available, these scales can be validated by comparing results to those obtained by using the “gold standard”.

Traditionally, noise in measures is treated as a function of how well it correctly categorises people, called classical measurement error.(11,12) But, an additional source of noise is also introduced by categorising observations on a continuous scale into groups and assuming a common value for all participants within each group: This additional variance is that between the true value of the observation and the value assigned to its group category, which we define here as the Variance From Categorisation (see Methods below). For example, the GAD-7 measures Generalised Anxiety Disorder (GAD) using 7, 4-level, questions which are summed into an 28-level scale. Each question asks participants to rate their symptoms over the past two weeks as lasting: not at all (scored 0), several days (scored as 1), over half the days (scored as 20, and nearly every day (scored as 3). If we assume that there is a continuous underlying liability to anxiety, then when this scale is used as either a continuous measure of Anxiety or transformed into a binary diagnostic proxy researchers implicitly assume a homogenous distribution of the participants’ actual liability to GAD within each measured group.(13)

Peto and colleagues concluded that large and simple randomised trials will need to answer some important questions robustly.(11,12) In order to facilitate the design of these studies, the aim of this study is to explore the relative merits of using simple to administer measures, such as a short questionnaire, which are cheap-but-noisy when compared to a gold standard. We assume that the two are similarly accurate, but differ in their precision. Specifically, we calculate the variance from categorising participants for uniform and normal distributions. We then use simulations and analytical methods to help determine when using a simple outcome measure will be more cost-effective than a gold standard.

## Methods

### Calculation of the Variance From Categorisation for different distributions

In our analysis we assume that a study ‘outcome’ can be measured perfectly on an interval or ratio scale comprising 100 levels (0 to 99). For example, if the outcome was subjective wellbeing, then the gold standard would classify patients between 0 (lowest possible subjective wellbeing) and 99 (highest possible wellbeing) with each integer between indicating increasingly favourable outcomes. When no measurement of outcome is made at all, every patient is treated equally and is assigned to the same outcome, equal to the midpoint of the outcome scale. This is equivalent to using an outcome rating scale with just one outcome category. For our analysis we assume that this scale measures outcome as 49.5 (the midpoint of the range of the possible values 0 to 99) for every participant. We additionally model scales with 2, 3, 5, 8 and 10 categories, and assume that the outcomes are the midpoints of the ranges of possible values covered by each category. For example, the measured outcomes using a scale with two outcome categories are the midpoints of the ranges of possible values covered by each category: 24.75 (midpoint of the range 0 to 49), and 74.5 (midpoint of the range 50 to 99) (see Figure 1 and Supplementary Table 1 for more detail).

**Figure 1:**
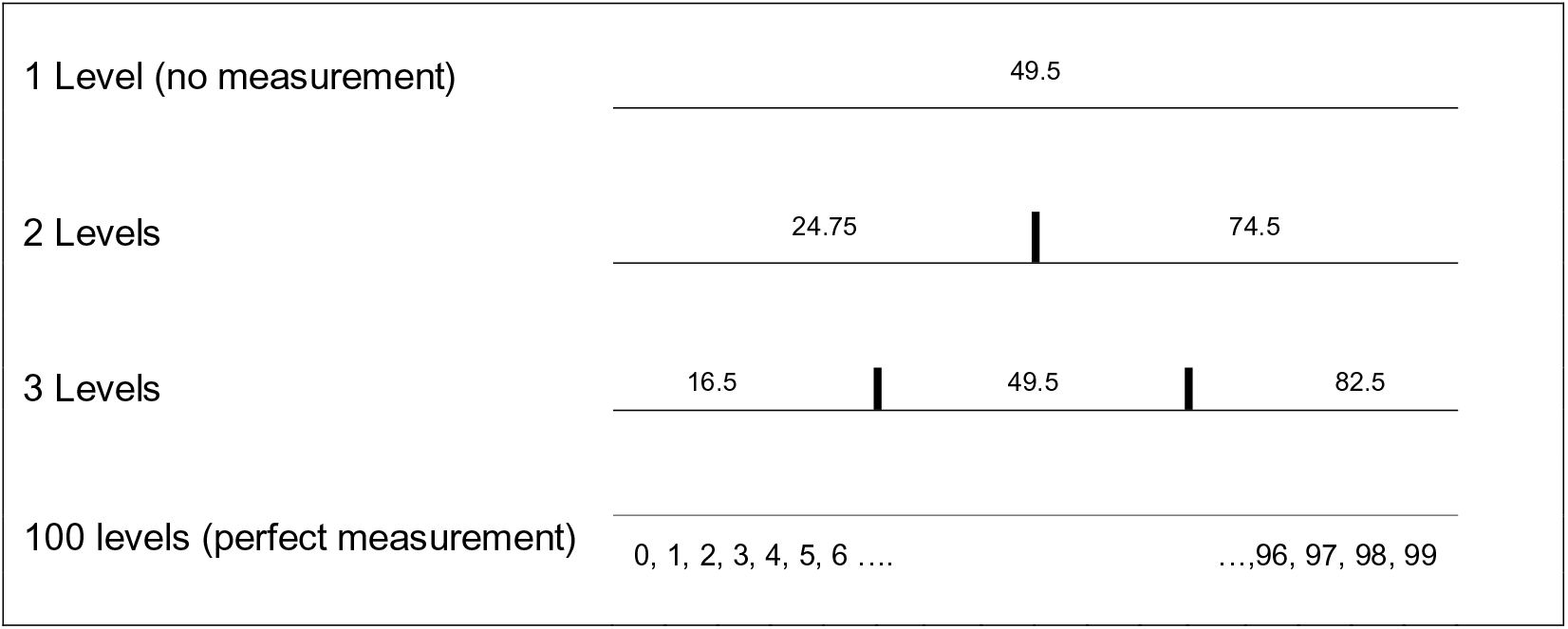
Outcome rating scales that subdivide the entire range of outcomes into 2, 3, .. outcome categories.

We define the variance due to the difference between the true value of the observation and the value assigned to its scale category (when a continuous scale is categorised) as the Variance From Categorisation (σ_c_^2^):

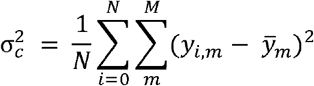

where *y*_*i,m*_ denotes the true outcome value in participant *i* within category *m* an 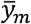 denotes the midpoint of the range of values in category *m. M* denotes the total number of categories and *N* the total number of participants.

We calculate the Variance from Categorisation for measures with 1, 2, 3, 5, 8, 10, or 15 outcome categories using a uniform distribution ranging from 0 to 99 and constrained discrete normal distributions with a range of 0 to 99, a mean of 49.5, and standard deviations ranging from 0.5 to 25 at 0.5 intervals. For the purpose of this study, we define no measurement as equivalent to having the population take a one category question which assigns everyone the mid-point value of the scale.

### Analytic conditions for simple measures to be cost effective

The sample to achieve a defined power in a parallel study design with two groups is given by the sample size formula: 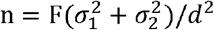, where n is the required sample size (assuming equal sized groups), F is a function of the critical values of the standard Normal distribution for a type I error of *α* and a type II error of *β, σ*_1_ and *σ*_2_ are the standard deviations in each group, and *d* is the difference in means to be detected. Using this formula and assuming that that there is no differential measurement error, we calculate the analytic conditions under which a simple measure will be more cost-effective than a more expensive but less noisy measure, where cost-effectiveness is defined as achieving the same power for a smaller cost.

### Measuring the effect of participant burden

To estimate the effect of increasing participant burden on response rates we re-analysed the data from two systematic reviews identified in a Google Scholar search on 9/9/21, Edwards et al (2009), and Rolstand et al (2011).(7,14) We used the data provided in the systematic reviews as a source of sample sizes and odds of responding in each arm of the randomised control trials in which participants were randomly assigned to either a long or short questionnaire. Where the reviews provided information on the nature of the long and short questionnaire this was used to calculate the ratio between the two. Else, BW extracted this information from the relevant studies included in the review. Additionally, studies which Rolstand et al noted as non-randomised were excluded. We then regressed the logged ratio of the odds of responding on the logged ratio of the length of questionnaires using the metareg function in Stata (V16).(15,16)

### Illustrative simulation of when simple measures are more cost-effective, and interactive web-app

Forming a definitive suggestion is difficult because many study specific factors will influence the equation on which design is more suitable. Such factors include the validity of the measures, the costs of both measures, the expected size of effect, and the amount of variability in the population. In addition, in practice, researchers generally do not measure measurement error using a variance statistic, but instead using correlation.

We therefore, additionally used a simulation to explore the comparative effects of using a cheap-but-noisy measure relative to a gold standard. In our simulation we model a power of 90%, an alpha of 5%, we additionally chose a low concurrent validity (r = 0.7) for the cheap-but-noisy measure, and a small effect size (mean difference = 0.1). Using parameters based on the CRASH-1 trial,(17) we model the cost of the gold standard as £50, and of the cheap-but-noisy measure as £5 per participant. We then measured the ratio of the costs of six measures (with 2, 3, 5, 8, 10, or 15 levels) for a range (0-25) of outcome standard deviations. In line with our previous simulations, we assumed the gold standard had 100 levels (0-99) and that that the outcome was normally distributed with a mean in the population of 49.5. Because the length of the gold standard was to some extent arbitrary, we ran the simulation with and without any effect of loss to follow up on response rates, additionally assuming that the cost of each lost participant was the same as the cost of a non-lost participant.

The simulations were produced in R,(18) and graphics were created using GG-Plot.(19) Because the parameters we chose may not be the ones of interest in practice, we also created an R-Shiny App, based off the above simulation, to allow researchers to use different parameters from the ones we have chosen.

## Results

### Variance from Categorization for uniform and normal distributions

The variance from categorization for the distributions defined above, using scales with 2, 3, 5, 8 and 10 categories, are shown in Table 1 and Supplementary Table 2. We found that most of the Variance from Categorisation was removed after adding 3 to 5 response scale categories. For example, for the uniform distribution the scale with 2 outcome categories resulted in a 75% reduction in variance compared to a single category measure taking the mid-point value. Increasing the number of outcome categories used in the scale to 3 or 5 resulted in an 89% and 96% reduction respectively, when compared to a single category taking the mid-point value. The reductions in measurement error achieved by using scales with larger numbers of outcome categories therefore diminished rapidly (Figure 2). For the normal distributions, the σ_c_^2^was also reduced by having a scale which was better calibrated to describing the variation in the data (demonstrated by the faster reduction in σ_c_^2^ as the standard deviation increased). The R code used to calculate the variance from categorisation can be found in Supplementary Table 3.

**Table 1:** Examples of variance induced by categorisation for different distributions. Each distribution has a range of 0-99, the normal distributions have a mean of 49.5.

| Number of categories | Variance from Categorizing (% reduction in compared to no measurement) |  |  |  |  |  |
| --- | --- | --- | --- | --- | --- | --- |
|  | Normal distribution, SD = 5 | Normal distribution, SD = 10 | Normal distribution, SD = 15 | Normal distribution, SD = 20 | Normal distribution, SD = 25 | Uniform distribution |
| 1 (no measurement) | 25.08 (0.0%) | 100.11 (0.0%) | 222.51 (0.0%) | 362.63 (0.0%) | 479.07 (0.0%) | 833.25 (0.0%) |
| 2 | 444.73 (-1623.8%) | 321.56 (-221.2%) | 247.66 (-11.3%) | 214.26 (40.9%) | 202.40 (57.8%) | 208.28 (75.0%) |
| 3 | 24.93 (3.4%) | 73.56 (26.5%) | 90.75 (59.21%) | 97.85 (70.0%) | 121.27 (74.7%) | 92.73 (88.8%) |
| 5 | 21.60 (16.3%) | 33.04 (67.0%) | 33.47 (85.0%) | 36.63 (89.9%) | 51.89 (89.2%) | 33.25 (96.0%) |
| 8 | 13.69 (46.9%) | 13.02 (87.0%) | 13.11 (94.1%) | 15.29 (95.8%) | 26.71 (94.4%) | 13.06 (98.4) |
| 10 | 8.41 (67.4%) | 8.33 (91.7%) | 8.40 (96.2%) | 10.29 (97.2%) | 20.54 (95.7%) | 8.25 (99.0%) |
| 15 | 3.74 (85.5%) | 4.30 (95.7%) | 5.26 (97.6%) | 5.61 (98.5%) | 5.67 (98.8%) | 5.46 (99.3%) |
| 100 (perfect measurement) | 0 (100%) | 0 (100%) | 0 (100%) | 0 (100%) | 0 (100%) | 0 (100.0%) |

**Figure 2:**
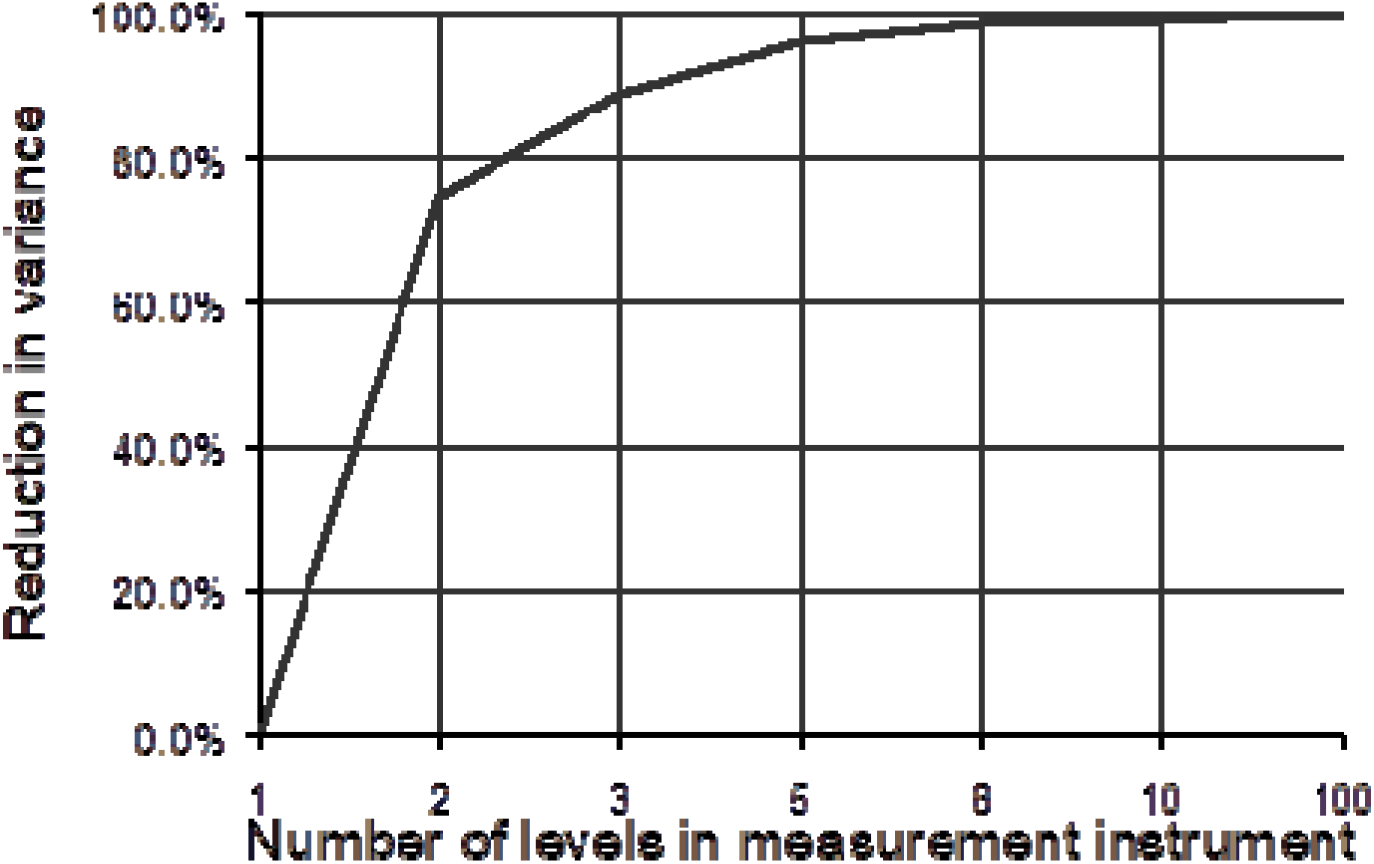
Percentage reduction in variance from categorisation from adding measurement levels, compared to no measurement.

### Analytic conditions under which simpler measures are more cost effective

The derivation of analytic conditions under which a simple measure is more cost effective than a noiseless but expensive measure, assuming equivalent measurement accuracy, can be found in Supplementary Table 4. This demonstrates that if the relative increase in cost from using the expensive measure is greater than the percentage increase in noise from using the simple measure, the simple measure will be more cost-effective.

### The impact of participant burden on non-response

Edwards et al (2009) included 56 studies in their meta-analysis of the impact on questionnaire length on final odds of responding, and Rolstand et al (2011) included a further 8 randomised controlled trails not included in Edwards et al. Of these, two had insufficient information in either the review or the study’s report to ascertain either the ratio or actual length of questionnaire in each arm. Of the remainder, 51 measured the length of questionnaire using the number of pages, 8 used the number of questions, 2 provided word counts, and 1 provided the time needed to complete the questionnaire (Supplementary Table 5).

The linear meta-regression is presented in Figure 4 and explained 24.8% of the variance in the odds of participants responding. This showed that for every increase of 1 in the log ratio of the length of questionnaire the log odds ratio for responding increased by −0.570 (95% CI −0.851 to −0.289, SE = 0.140, p < 0.001). There was not, additionally, evidence that the intercept was different from zero (beta = 0.103, SE = 0.155, p = 0.509). We additionally did not find any evidence that adding in a quadratic term improved model fit (new r^2^ = 23.42%). Converting these parameters from the log-log scale to the natural scale, we therefore find a power law relationship: the ratio in OR = RQ^-0.57^, where OR is the ratio in the odds of responding to the long questionnaire relative to the short questionnaire, and RQ is the ratio in questionnaire length of long questionnaire to the short questionnaire. This implies that doubling the length of a questionnaire would reduce the response rate by around 65%, and therefore require asking around 50% more people to participate to achieve the same number of outcome observations (Figure 5).

**Figure 3a:**
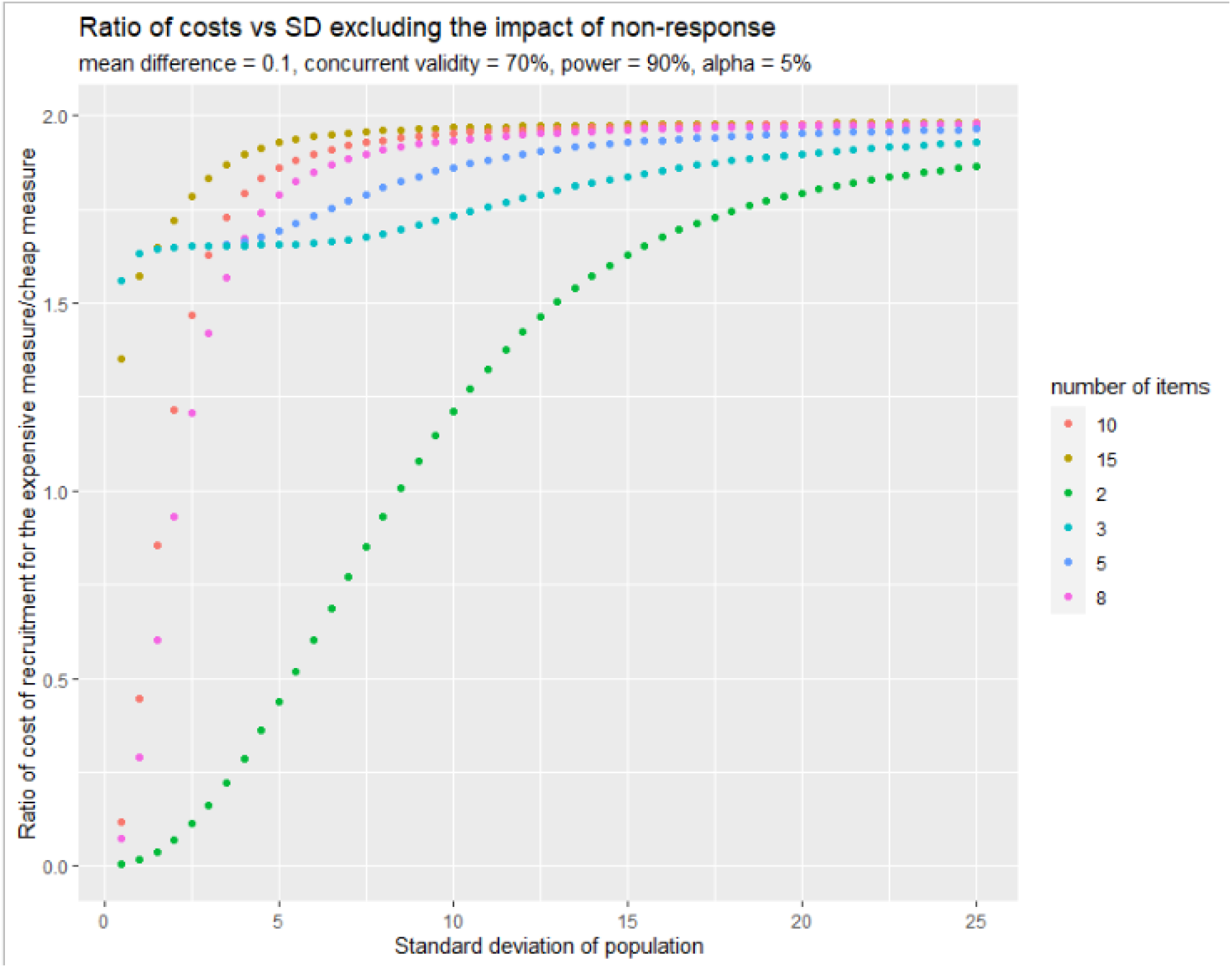
Illustrative results of simulation, excluding the impact of non-response.

**Figure 3b:**
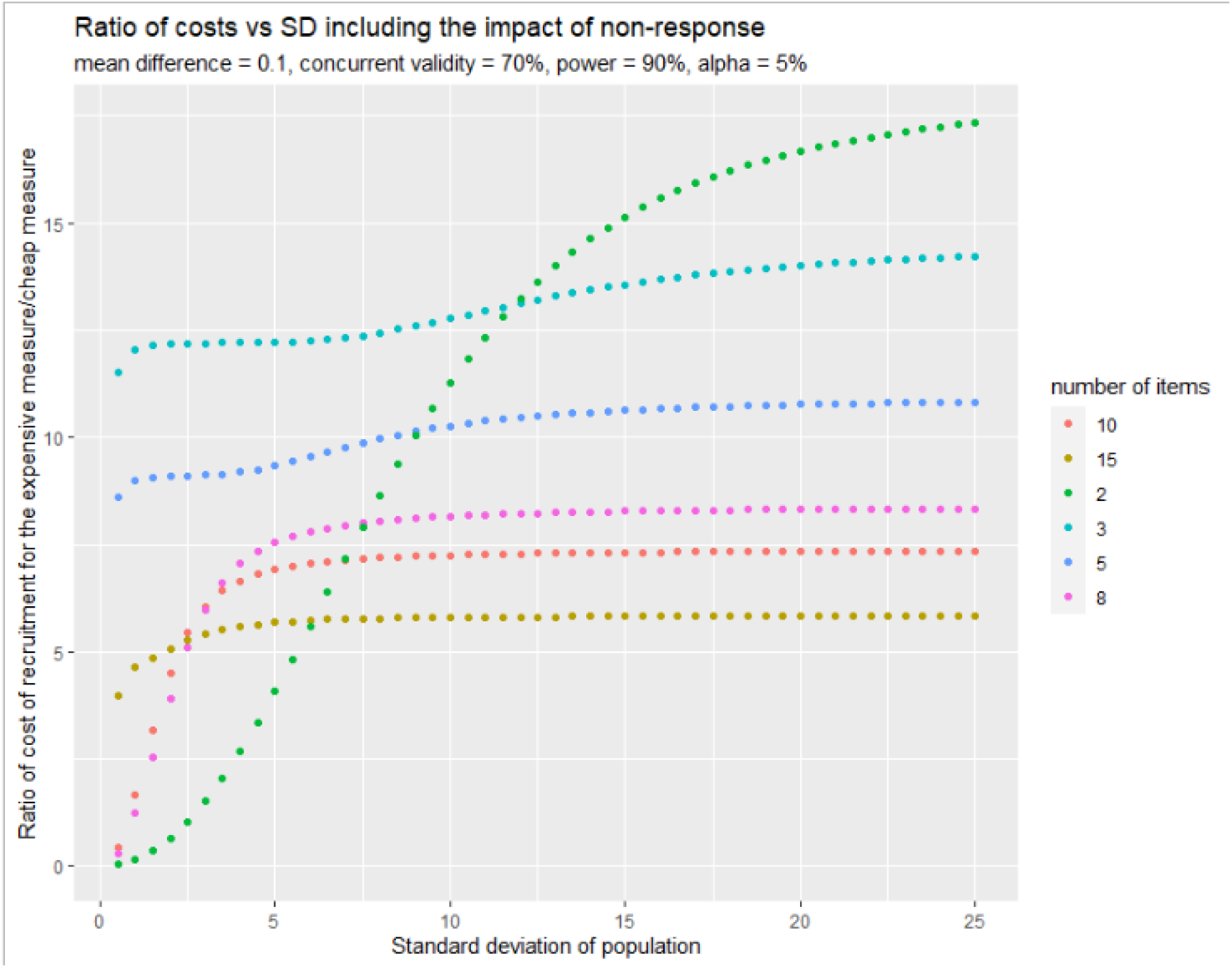
Illustrative results of simulation, including the impact of non-response.

**Figure 4:**
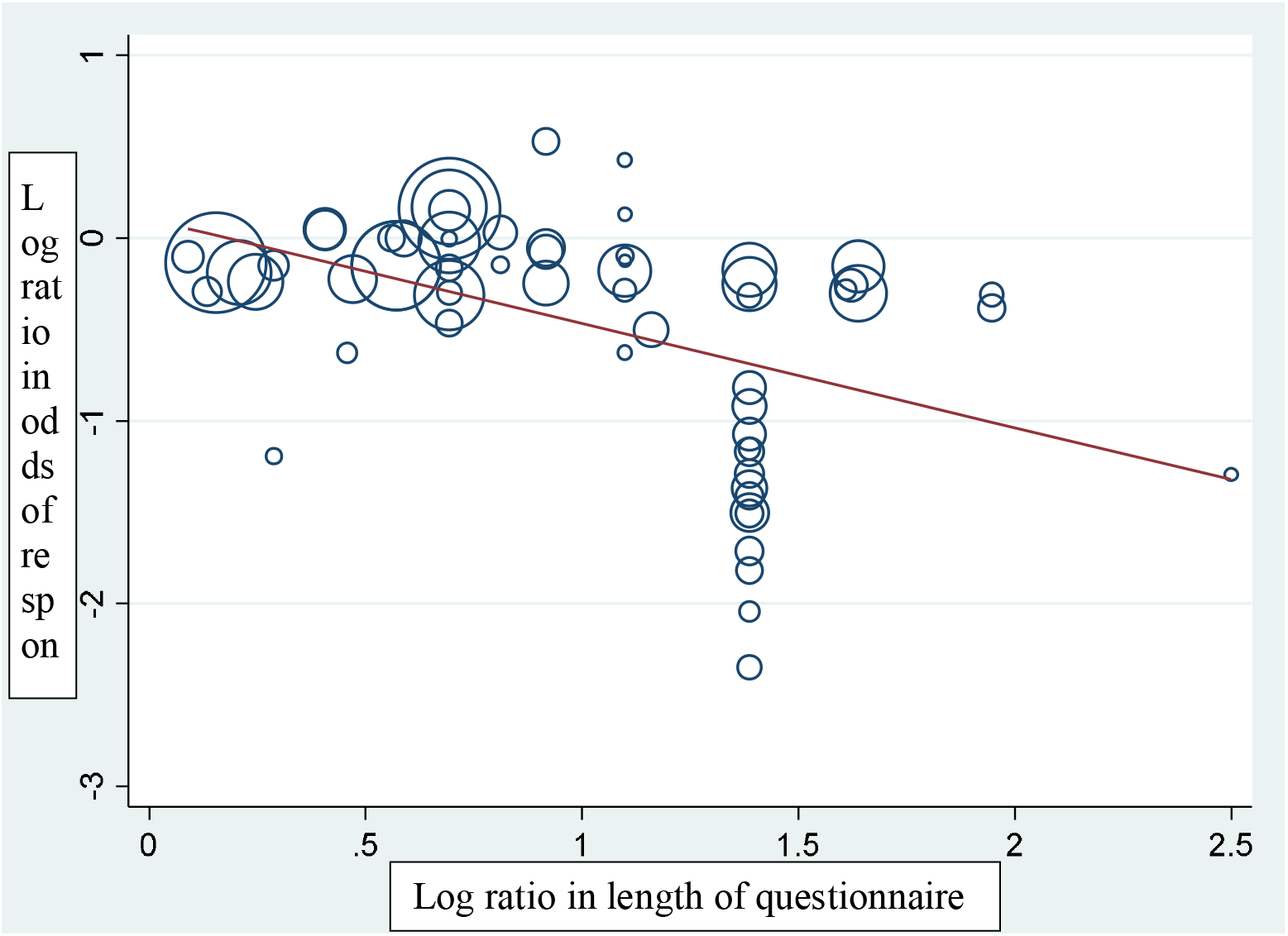
Results of the meta-regression, with both variables presented on the log-log scale. Both ratios are defined as the ratio in length of long questionnaire to short questionnaire.

**Figure 5:**
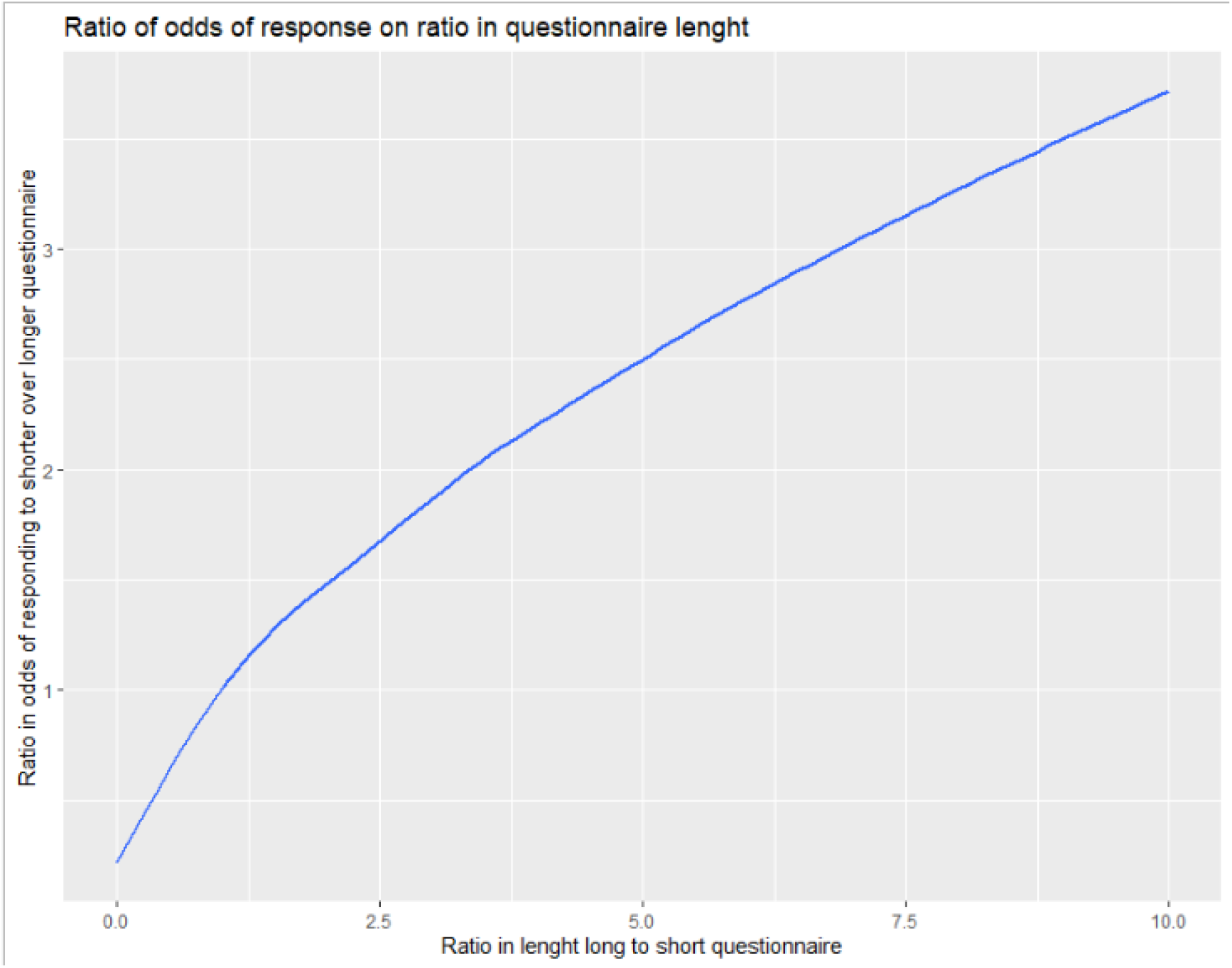
Illustration of the effect of increasing questionnaire length on reducing response rates from meta-regression. The y-axis can be interpreted as the multiple of how many more participants would be needed to achieve the same number of responses given how many times larger the questionnaire being used is.

### Simulation and interactive web-app

Our simulation found that most cheap-but-noisy measures outperformed the gold standard when the standard deviation in the population was approximately greater than 1 (Figure 3). Ignoring the effect of participant burden means that the longer questionnaires, with a lower variance of categorisation, performed best (Figure 3a). However, including an effect for loss to follow up resulting in shorter questionnaires outperforming longer ones (Figure 3b). We additionally created an R-shiny app (available at https://benjiwoolf.shinyapps.io/cheapbutnoisymeasures/) to allow readers to further explore our simulation using different parameters. The R code used in the simulation is available in Supplementary Table 3.

## Discussion

### Principle findings

We have shown that cheap-but-noisy measures, such as short questionnaires, may be more cost-effective than their gold standard counterparts. Specifically, a cheap-but-noisy measure will be more cost-effective than a gold standard measure with no measurement error or Variance from Categorisation if the relative increase in cost is greater than the relative increase in noise from measurement error and the Variance from Categorisation, providing the cheap-but-noisy measure maintains similar accuracy of the gold standard. This could be achieved by using well-validated short form questionnaires, and we would encourage further development and validation of such measures for a wide range of clinical outcomes.

We introduced the notion of Variance from Categorisation 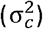 and showed that even a very simple measure, such as five yes/no questions or one five level Likert scale, eliminates the vast majority of the 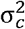 compared to functional no-measurement. 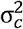 was also reduced for normally distributed variables when the scale was calibrated to the variance expected within the population, with more homogeneous populations needing more sensitive scales. This demonstrates the importance of using a measure calibrated to the study population.

Our simulation demonstrates that the utility of a simple questionnaire, with few items or response categories, is inversely related to the required sensitivity of the test. If the measure is unable to detect effect sizes as large (or small) as they are expected to be, then the cheap measure is guaranteed to be less useful than a more sensitive measure, even if the gold standard is substantially more expensive. This is analogous to the final part of Smeden et al.’s Triple Whammy of Measurement error, that it can mask features of the data such as effect modification and non-linear associations.(20) Although this is an unsurprising finding, it raises an important caveat that the most appropriate measure will vary depending on factors unique to every study, and that our results do not warrant the use of cheap-but-noisy measures in every circumstance.

For many studies there is a cost associated with non-response. For example, when there is a sunk cost due to the allocation process/cost of the intervention, such as a pharmaceutical trial having to cover the cost of the active and placebo drugs taken by participant. Even if the intervention is free, an outcome measured through a postal questionnaire, e.g. in psychiatric trial, will have a cost associated with posting and printing unreturned questionnaires. An important implication of the association between participant burden (e.g. questionnaire length) and non-response is that, in studies with a cost of non-response, the analytic solution for when a simple measure is more cost-effect, will under-estimate the saving associated with using a simpler measure because the expected higher response rate. In a meta-regression of a previously conducted Cochrane systematic review we estimate that the ratio of response rates has an inverse power law relationship with questionnaire length, such that doubling the participant burden (measured as questionnaire size) will reduce the response rate by around one third.

One important source of systematic bias is differential response (selection bias). As this study shows, simpler questionnaires have meaningfully lower loss to follow-up. This implies that an additional advantage of the use of cheap-but-noisy measures is a reduction in the risk of selection bias. For example, the International Stroke Trial assessed disability after stroke for 19435 participants.[7] Conventional outcome measures (e.g. the Barthel Index and Oxford Handicap Scale) were considered to be too complicated and expensive. Instead, two simple questions with a reasonable validity relative to the Barthel Index and the Oxford Handicap Scale were used to measure handicap.[17,18] This allowed participants to be classified into three levels: needing help (‘dependent’), not needing help but still with some handicap (‘independent’), and those not needing help and with no handicap (‘independent and recovered’). At follow up, six months after randomisation, the trial achieved a 99% response rate, and evidence of a clinically important treatment effect. Even if all data was missing not at random, the potential for serious selection bias is minimal. It seems likely that this high response rate may have been influenced by the decision to use a simple outcome measure.

### Strengths and limitations of the study

We did not consider two other types of non-differential measurement error: calibration error (which occurs when assigning incorrect units to a scale, for example using a yard stick to measure a metre without any unit transformations) and non-differential bias (which occurs when a constant number of units is added to all readings from a scale, e.g. parallax error).(21) Although both of these can invalidate results, psychometric-type questionnaires are typically standardised, and both of these errors are removed by standardising the outcome and calculating the standardised mean difference. Therefore, a caveat of our simulation is that we assume the scales are standardised.

The meta-regression should, additionally, be taken with a grain of salt. Firstly, neither review explored risk of bias, which is known to lead to an inflation of estimates.(3) The results may also not generalise to a setting relevant for our simulation. 82% of studies used the number of pages as their measure of questionnaire length, while our simulation explored the impact of varying the number of questions. The relationship between the number of questions and pages may not always be linear: two short questionnaires may both be one page but if the first is only half a page long the second may be almost twice as long while still being the same number of pages. Likewise, Edwards et al.’s review focused on postal questionnaires, and the point estimate may not generalise to electronic questionnaires. For example, we are aware of a more recent controlled trial exploring the effect of questionnaire length in electronic questionnaires which did not find evidence of a negative impact of longer questionnaires.(22) Finally, our meta-regression deviated from systematic review best-practices because additional information was extracted by only one author.

The parameters of the simulation obviously limit the generalisability of its results. Firstly, we only simulated a limited set of parameters which may not generalise. For example, it is unlikely that a large proportion of potential scales will have the same range and mean as the one we simulated. Because of this we created an R-shiny app (available at https://benjiwoolf.shinyapps.io/cheapbutnoisymeasures/) to allow researchers to use alternative parameters. Finally, it is worth noting that we assumed a certain level of validity in the cheap-but-noisy measures, and that our results do not warrant the use of completely unvalidated measures. We would therefore suggest that authors explore the use of validated short form questionnaires before constructing an unvalidated outcome measure.

Finally, that we assume a univariate analysis. Although this is appropriate for some study designs like randomised controlled trails and genome wide analysis studies, these results may not be applicable in other settings. Importantly, in a multivariate analysis, for example when adjusting for confounders or in a mediation analysis, non-differential measurement error can create a bias in either direction.(20)

### Selection bias - Information Bias trade off

Our study has only explored the benefits of simple measures in terms of the power of a clinical trial. However, one important limitation is that simple measures may increase risk of (information) bias due to differential measurement error. Differential measurement error occurs when the error is not random but depends on some other factor related to the pertinent.(23,24) Risk of bias is arguably more important than improving power because random error can be eliminated in a systematic review and meta-analysis of many small studies, while residual bias is not. Simple measures, like a questionnaire, may be more suspectable to bias than a more expensive measure for two reasons.

Firstly, a simple questionnaire can be influenced by social and psychological factors which a more expensive objective measure, like a biometric reading, will not be. For example, a researcher may subtitle change the way they ask a question about alcohol consumption based of their perception of a participant (‘interviewer bias’), while a participant’s knowledge of social expectations may lead them to downplay how much alcohol they have drunk (‘social desirability bias’). On the other hand, neither the researcher’s nor participant’s expectations will influence a breathalyser reading. A well-designed questionnaire should be able to reduce this type of information bias. For example, psychometricians can use methods such as control questions, reverse coding, and an independent-rater to reduce the impact of these biases on study quality. A limitation of these methods is that they increase the complexity of outcome measures, and therefore may undermine both the simplicity and cheapness of the cheap-but-noisy measure. This type of bias can also be attenuated by improving the study design. Blinding can be used to reduce the possibility that any bias is differential across exposure status. Likewise, the use of anonymised postal or on-line questionnaires may reduce perceived social pressures or other (interviewer) bias due to having study personnel requesting information from participants in person.

A second source of differential measurement error could come from the measure itself. Many outcomes are intrinsically complex or multi-dimensional, obvious examples being socio-economic position or frailty. A risk of using a simple measure is that it may not capture all of the desired dimensions of the outcome of interest. When this occurs it is likely that the more simple measure will produce incorrect estimates of effect. For example, frailty is often thought to involve both physical and psychological dimensions.(25) An evaluation of an intervention designed to reduce frailty may produce misleading results if it only measures the psychological impact of frailty, or potentially miss the entire effect if the intervention’s impact is mostly mediated by reducing physical frailty.

Because these sources of information bias are intrinsic to the simplification process they are a limiting factor on the utility of cheap-but-simple measures. The higher participant burden in most gold-standard measures, however, increases risk of selection bias in studies that use them when compared to a cheap-but-noisy alternative. This implies the existence of a second quality-quantity trade off not explored in this simulation. The amount of selection and information bias in a study will vary depending on each study’s methods, and, if measurable, can only be quantified post-hoc. It is therefore impossible to provide universally applicable methods prescriptions beyond attempting to minimise the overall risk of bias in a study. With this in mind, we believe that cheap and noisy measures should be considered with caution in studies where information bias is likely, but may be useful in reducing the overall risk of bias in studies with a greater risk of selection bias than information bias.

### Unanswered questions and future research

There are many outcomes which are continuous but may be easier to measure or model using a binary or categorical outcome. For example, recovery form a stroke is in theory a continuous outcome varying from no recovery (e.g. 0) to full recovery (e.g. 100). However, it has been argued that recovery can be efficiently measured using a four level ordinal outcome measured using two yes/no questions.(26–28) The practice of dichotomising continuous outcomes remains controversial because cut points are often arbitrary and generally reduce power.(29) One application of Variance From Categorisation may be as a conceptual aid in this discussion. It could be argued that, given justifiable thresholds, categorisation may reduce the total error when the Variance From Categorisation is less than the sum of the squared residuals from using a continuous parametric model for the outcome. As an extreme example, power law distributed variables with a negative exponent less than three, such as the supposedly common Pareto 80:20 distribution,(30,31) will not have a finite variance. Because they fail to meet a condition for the central limit theorem, the error in the estimation of their mean cannot be assumed to be normally distributed. Although beyond the scope of this paper, it is plausible that in such instances the total error in the statistical model for the outcome could be reduced by categorising it.

Finally, classical measurement error in the exposure of a bivariate analysis, such as a linear regression model with one regressor, will generally lead to regression dilution bias in which the effect estimate is biased toward the null. Various sensitivity analysis, such as regression calibration and SIMEX, have been developed to attenuate this bias.(32) To the extent to which a cheap-but-noisy measure contains more classical error, using these methods seems sensible. However, the extent to which these methods may be useful in overcoming any bias due to Variance from Categorisation remains unclear.

### Conclusion and implications for outcome measures in clinical trials

Clinical trials will become more cost effective by employing cheap-but-noisy outcome measures, such as a simple questionnaire, when the relative increase in cost between the cheap-but-noisy measure and its alternative is greater than the relative increase in noise, assuming no bias. Simple questionnaires, with a given level of validity, have the added advantages of reducing loss to follow-up by improving response rates and not adding large amounts of noise. However, the relative merits of doing so will vary from study to study. Importantly, any increase in power and reduction of susceptibility to selection bias must be balanced against a potential increase in information bias. Box 1 provides a checklist of questions we hope will provide readers with a useful screen for when not to use a cheap-but-noisy measure. Finally, we have assumed throughout the existence of a previously created, and validated, questionnaire that could be used as a cheap-but-noisy outcome measure. Although questionnaires are becoming increasingly popular as health measures, see for example reference (33–36), we would encourage the development of a wider range of questionnaires to enable their use as end points in clinical trials.

#### Box 1

Checklist for screening cheap-but-noisy measures

1. Is there a candidate cheap but noisy measure?
2. What are the units of the measure? If it is not standardised, is there a risk of calibration error or a non-differential bias like parallax error?
3. Does the cheap-but-noisy measure have sufficient sensitivity to detect the expected effect and not mask any important variation or features of the data?
4. Is the outcome simple or multidimensional? If multidimensional, does the cheap-but-noisy measure capture signal from all relevant dimensions?
5. Is there a material risk of response biases like interviewer or recall bias?
6. Are there changes to the measure design (such as control questions, reverse coding, independent-rater, etc.) or study design (e.g. blinding of participants and study personnel, online questionnaires, etc.) that could attenuate a response bias?
7. Biased off question 4. to 6. what is the likely overall size and direction of any information bias that using a cheap-but-noisy measure could introduce?
8. Is there an expected increase in sample size from using a cheap-but-noisy measure? What is the expected size and direction of a reduction in risk of selection bias?
9. Does the reduction in risk of selection bias outweigh any increase in the risk of information bias?
10. Has a cheap-but-noisy measure been validated? If not, we should suggest authors conduct a validation study if possible.
11. How much less expensive is the Is the cheap-but-noisy measure?
12. How much more noisy is the cheap-but-noisy measure?
13. Howe reliable is the estimation of the above two numbers?
14. Is the cheap-but-noisy measure cost effective?

## Data Availability

All data produced in the present work are contained in the manuscript

## Author contributions

PE defined σ_c_^2^. BW designed, implemented the study. BW and PE wrote up the report. HRB confirmed the analytic solution. HP created the R shiny app. All authors provided feedback on the manuscript.

## References

1. Yusuf S, Collins R, Peto R. Why do we need some large, simple randomized trials? Stat Med. 1984 Dec;3(4):409–22.

2. Peto R, Collins R, Gray R. Large-scale randomized evidence: large, simple trials and overviews of trials. J Clin Epidemiol. 1995 Jan;48(1):23–40.

3. Higgins JP, Savović J, Page MJ, Elbers RG, Sterne JA. Assessing risk of bias in a randomized trial. In: Cochrane Handbook for Systematic Reviews of Interventions [Internet]. John Wiley & Sons, Ltd; 2019 [cited 2022 May 16]. p. 205–28. Available from: https://onlinelibrary.wiley.com/doi/abs/10.1002/9781119536604.ch8

4. Devine OJ, Smith JM. Estimating sample size for epidemiologic studies: the impact of ignoring exposure measurement uncertainty. Stat Med. 1998 Jun 30;17(12):1375–89.

5. Edwards P. Questionnaires in clinical trials: guidelines for optimal design and administration. Trials. 2010 Jan 11;11(1):2.

6. Hajian-Tilaki K. Sample size estimation in epidemiologic studies. Caspian J Intern Med. 2011;2(4):289–98.

7. Edwards P, Roberts I, Clarke M, DiGuiseppi C, Pratap S, Wentz R, et al. Methods to increase response rates to postal questionnaires. Cochrane Database Syst Rev. 2007 Apr 18;(2):MR000008.

8. Likert R. A technique for the measurement of attitudes. Archives of Psychology. 1932;22 140:55–55.

9. Stevens SS. On the Theory of Scales of Measurement. Science. 1946;103(2684):677–80.

10. Plomin R, Haworth CMA, Davis OSP. Common disorders are quantitative traits. Nat Rev Genet. 2009 Dec;10(12):872–8.

11. Streiner DL. Health Measurement Scales: A practical guide to their development and use. 4th edition. Oxfordlll; New York: Oxford University Press, USA; 2008. 450 p.

12. Modern Psychometrics (International Library of Psychology): Amazon.co.uk: Rust, John: 9780415203418: Books [Internet]. [cited 2022 May 16]. Available from: https://www.amazon.co.uk/Modern-Psychometrics-Psychological-Assessment-International/dp/0415203414/ref=sr_1_3?crid=29IA0CMPVDRH7&keywords=modern+psychometrics&qid=1652697321&s=books&sprefix=modern+psychometrics%2Cstripbooks%2C68&sr=1-3

13. Williams N. The GAD-7 questionnaire. Occupational Medicine. 2014 Apr 1;64(3):224.

14. Rolstad S, Adler J, Rydén A. Response Burden and Questionnaire Length: Is Shorter Better? A Review and Meta-analysis. Value in Health. 2011 Dec 1;14(8):1101–8.

15. Harbord R, Higgins J. METAREG: Stata module to perform meta-analysis regression [Internet]. 2009 [cited 2022 May 16]. (Statistical Software Components). Available from: https://econpapers.repec.org/software/bocbocode/S446201.htm

16. Gutierrez RG. Stata. WIREs Computational Statistics. 2010;2(6):728–33.

17. Edwards P, Arango M, Balica L, Cottingham R, El-Sayed H, Farrell B, et al. Final results of MRC CRASH, a randomised placebo-controlled trial of intravenous corticosteroid in adults with head injury-outcomes at 6 months. Lancet. 2005 Jun 4;365(9475):1957–9.

18. R Core Team. R: A language and environment for statistical computing. R Foundation for Statistical Computin [Internet]. 2021. Available from: https://www.R-project.org/

19. Wickham H. ggplot2. WIREs Computational Statistics. 2011;3(2):180–5.

20. van Smeden M, Lash TL, Groenwold RHH. Reflection on modern methods: five myths about measurement error in epidemiological research. International Journal of Epidemiology. 2020 Feb 1;49(1):338–47.

21. Pierce BL, VanderWeele TJ. The effect of non-differential measurement error on bias, precision and power in Mendelian randomization studies. International Journal of Epidemiology. 2012 Oct 1;41(5):1383–93.

22. Blumenberg C, Menezes AMB, Gonçalves H, Assunção MCF, Wehrmeister FC, Barros FC, et al. The role of questionnaire length and reminders frequency on response rates to a web-based epidemiologic study: a randomised trial. International Journal of Social Research Methodology [Internet]. 2019 Jun 17 [cited 2022 May 16]; Available from: https://www.tandfonline.com/doi/full/10.1080/13645579.2019.1629755

23. Davey Smith G, Hemani G. Mendelian randomization: genetic anchors for causal inference in epidemiological studies. Human Molecular Genetics. 2014 Sep 15;23(R1):R89–98.

24. Hernán MA, Robins JM. Causal Inference: What If. :311.

25. Gobbens RJJ, Assen MALM van, Luijkx KG, Wijnen-Sponselee MT, Schols JMGA. Determinants of Frailty. Journal of the American Medical Directors Association. 2010 Jun 1;11(5):356–64.

26. Lindley RI, Waddell F, Livingstone M, Sandercock P, Dennis MS, Slattery J, et al. Can Simple Questions Assess Outcome after Stroke? CED. 1994;4(4):314–24.

27. Dorman P, Dennis M, Sandercock P. Are the modified “simple questions” a valid and reliable measure of health related quality of life after stroke? United Kingdom Collaborators in the International Stroke Trial. J Neurol Neurosurg Psychiatry. 2000 Oct;69(4):487–93.

28. McKevitt C, Dundas R, Wolfe C. Two Simple Questions to Assess Outcome After Stroke. Stroke. 2001 Mar;32(3):681–6.

29. Categorizing Continuous Variables - research methods / measurement [Internet]. Datamethods Discussion Forum. 2022 [cited 2022 May 16]. Available from: https://discourse.datamethods.org/t/categorizing-continuous-variables/3402

30. Micceri T. The unicorn, the normal curve, and other improbable creatures. 1989;

31. Newman MEJ. Power laws, Pareto distributions and Zipf’s law. Contemporary Physics. 2005 Sep;46(5):323–51.

32. He W, Xiong J, Yi GY. SIMEX R Package for Accelerated Failure Time Models with Covariate Measurement Error. Journal of Statistical Software. 2012 Jan 25;46:1–14.

33. Spinou A, Siegert RJ, Guan WJ, Patel AS, Gosker HR, Lee KK, et al. The development and validation of the Bronchiectasis Health Questionnaire. Eur Respir J. 2017 May;49(5):1601532.

34. Jenkinson C, Stewart-Brown S, Petersen S, Paice C. Assessment of the SF-36 version 2 in the United Kingdom. Journal of Epidemiology & Community Health. 1999 Jan 1;53(1):46– 50.

35. Harwood RH, Rogers A, Dickinson E, Ebrahim S. Measuring handicap: the London Handicap Scale, a new outcome measure for chronic disease. BMJ Quality & Safety. 1994 Mar 1;3(1):11–6.

36. Harwood RH, Ebrahim S. The validity, reliability and responsiveness of the Nottingham Extended Activities of Daily Living scale in patients undergoing total hip replacement. Disability and Rehabilitation. 2002 Jan 1;24(7):371–7.

