## Supplementary Table 3 for "Silence is golden, by my measures still see: why cheap-but-noisy outcome measures can be more cost effective than gold standards"

##############################################################################################################
##############################################################################################################
###############################excel spreadsheet in R ######################################################
##############################################################################################################
##############################################################################################################

####################################
# installing packages 
###############################

install.packages('tidyverse')
install.packages("truncnorm")
install.packages('gtools')

#install.packages 
library(gtools)
library(ggplot2)
library(tidyverse)
library(truncnorm)


##############################################################################################################
#############################################################################################################
# calculating the variance of categorization 
##############################################################################################################
#############################################################################################################


# calculating the variance of categorization with a uniform distribution 
#############################################################################

df<-1
df<-data.frame(df)

#creating the data frame
n100<-c(0:99)
vom<- data.frame(n100)


# calculate variance of categorization for 1 level
vom$c1<-49.5
vom$vom1<-(vom$n100-vom$c1)^2
df$vom1<-mean(vom$vom1)

# calculate variance of categorization for 2 levels
vom$c2<-74.5
vom$c2[vom$n100<50]<-24.75
vom$vom2<-(vom$n100-vom$c2)^2
df$vom2<-mean(vom$vom2)

# calculate variance of categorization for 3 levels
vom$c3<-82.5
vom$c3[vom$n100<65]<-49.5
vom$c3[vom$n100<34]<-16.5
vom$vom3<-(vom$n100-vom$c3)^2
df$vom3<-mean(vom$vom3)

# calculate variance of categorization for 5 levels
vom$c5<-89.5
vom$c5[vom$n100<80]<-69.5
vom$c5[vom$n100<60]<-49.5
vom$c5[vom$n100<40]<-29.5
vom$c5[vom$n100<20]<-9.5
vom$vom5<-(vom$n100-vom$c5)^2
df$vom5<-mean(vom$vom5)

# calculate variance of categorization for 8 levels
vom$c8<-93.5
vom$c8[vom$n100<88]<-81
vom$c8[vom$n100<75]<-68.5
vom$c8[vom$n100<63]<-56
vom$c8[vom$n100<50]<-43.5
vom$c8[vom$n100<38]<-31
vom$c8[vom$n100<25]<-18.5
vom$c8[vom$n100<13]<-6
vom$vom8<-(vom$n100-vom$c8)^2
df$vom8<-mean(vom$vom8)

# calculate variance of categorization for 10 levels
vom$c10<-94.5
vom$c10[vom$n100<90]<-84.5
vom$c10[vom$n100<80]<-74.5
vom$c10[vom$n100<70]<-64.5
vom$c10[vom$n100<60]<-54.5
vom$c10[vom$n100<50]<-44.5
vom$c10[vom$n100<40]<-34.5
vom$c10[vom$n100<30]<-24.5
vom$c10[vom$n100<20]<-14.5
vom$c10[vom$n100<10]<-4.5
vom$vom10<-(vom$n100-vom$c10)^2
df$vom10<-mean(vom$vom10)

# calculate variance of categorization for 15 levels
vom$c15<-96
vom$c15[vom$n100<93]<-89
vom$c15[vom$n100<86]<-82.5
vom$c15[vom$n100<80]<-76
vom$c15[vom$n100<73]<-69
vom$c15[vom$n100<66]<-62.5
vom$c15[vom$n100<60]<-56
vom$c15[vom$n100<53]<-49
vom$c15[vom$n100<46]<-42.5
vom$c15[vom$n100<40]<-36
vom$c15[vom$n100<33]<-34
vom$c15[vom$n100<26]<-22.5
vom$c15[vom$n100<19]<-16
vom$c15[vom$n100<13]<-9
vom$c15[vom$n100<6]<-2.5
vom$vom15<-(vom$n100-vom$c15)^2
df$vom15<-mean(vom$vom15)

setwd("C:/Users/nf18217/OneDrive - University of Bristol/Documents/Projects/active/phil ME")
write.csv(df,'uniform voc.csv')


# calculating the variance of categorization with a normal distribution 
#############################################################################

# Defining the data frame. 
df<- c(1:50)
df<- data.frame(df)

# Range of standard deviations to be considered 
df$sd<- seq(0.5, 25, by=0.5) 

df$vom100<-0 # variance of measurement with gold standard 
df$vom15<-5.46 # arbitrary values used to create the variables in the data set. 
df$vom10<- 8.25 # arbitrary values used to create the variables in the data set. 
df$vom8<- 13.06 # arbitrary values used to create the variables in the data set. 
df$vom5<- 33.25 # arbitrary values used to create the variables in the data set. 
df$vom3<-92.73 # arbitrary values used to create the variables in the data set. 
df$vom2<-208.28 # arbitrary values used to create the variables in the data set. 
df$vom1<-833.25 # arbitrary values used to create the variables in the data set. 

vom<-c(1:100000000)  # size of data set used to calculate the VOM. 
vom<-data.frame(vom)

#Loop for calculating the size VOM
for (i in df$sd) {
  # create simulated data set
  vom$n<-rtruncnorm(100000000, a=0, b=99, mean=49.5, sd=i) 
  # round to whole numbers
  vom$n100<-round(vom$n, digits = 0)
  
  # calculate variance of categorization for 1 level
  vom$c1<-49.5
  vom$vom1<-(vom$n100-vom$c1)^2
  df$vom1[df$sd==i]<-mean(vom$vom1)
  
  # calculate variance of categorization for 2 levels
  vom$c2<-74.5
  vom$c2[vom$n100<50]<-24.75
  vom$vom2<-(vom$n100-vom$c2)^2
  df$vom2[df$sd==i]<-mean(vom$vom2)
  
  # calculate variance of categorization for 3 levels
  vom$c3<-82.5
  vom$c3[vom$n100<65]<-49.5
  vom$c3[vom$n100<34]<-16.5
  vom$vom3<-(vom$n100-vom$c3)^2
  df$vom3[df$sd==i]<-mean(vom$vom3)

  # calculate variance of categorization for 5 levels
  vom$c5<-89.5
  vom$c5[vom$n100<80]<-69.5
  vom$c5[vom$n100<60]<-49.5
  vom$c5[vom$n100<40]<-29.5
  vom$c5[vom$n100<20]<-9.5
  vom$vom5<-(vom$n100-vom$c5)^2
  df$vom5[df$sd==i]<-mean(vom$vom5)
  
  # calculate variance of categorization for 8 levels
  vom$c8<-93.5
  vom$c8[vom$n100<88]<-81
  vom$c8[vom$n100<75]<-68.5
  vom$c8[vom$n100<63]<-56
  vom$c8[vom$n100<50]<-43.5
  vom$c8[vom$n100<38]<-31
  vom$c8[vom$n100<25]<-18.5
  vom$c8[vom$n100<13]<-6
  vom$vom8<-(vom$n100-vom$c8)^2
  df$vom8[df$sd==i]<-mean(vom$vom8)
  
  # calculate variance of categorization for 10 levels
  vom$c10<-94.5
  vom$c10[vom$n100<90]<-84.5
  vom$c10[vom$n100<80]<-74.5
  vom$c10[vom$n100<70]<-64.5
  vom$c10[vom$n100<60]<-54.5
  vom$c10[vom$n100<50]<-44.5
  vom$c10[vom$n100<40]<-34.5
  vom$c10[vom$n100<30]<-24.5
  vom$c10[vom$n100<20]<-14.5
  vom$c10[vom$n100<10]<-4.5
  vom$vom10<-(vom$n100-vom$c10)^2
  df$vom10[df$sd==i]<-mean(vom$vom10)

  # calculate variance of categorization for 15 levels
  vom$c15<-96
  vom$c15[vom$n100<93]<-89
  vom$c15[vom$n100<86]<-82.5
  vom$c15[vom$n100<80]<-76
  vom$c15[vom$n100<73]<-69
  vom$c15[vom$n100<66]<-62.5
  vom$c15[vom$n100<60]<-56
  vom$c15[vom$n100<53]<-49
  vom$c15[vom$n100<46]<-42.5
  vom$c15[vom$n100<40]<-36
  vom$c15[vom$n100<33]<-34
  vom$c15[vom$n100<26]<-22.5
  vom$c15[vom$n100<19]<-16
  vom$c15[vom$n100<13]<-9
  vom$c15[vom$n100<6]<-2.5
  vom$vom15<-(vom$n100-vom$c15)^2
  df$vom15[df$sd==i]<-mean(vom$vom15)
  
  }


#standard error for the values used in the VOM calculation
df$vomse<-df$sd/100000000^0.5

setwd("C:/Users/nf18217/OneDrive - University of Bristol/Documents/Projects/active/phil ME")
write.csv(df,'normal voc.csv')


##############################################################################################################
##############################################################################################################
############################## visualizing the impact of participant burden ###################################
##############################################################################################################
##############################################################################################################

# ln odds ratio for responding = 0.6*ln ratio size 

leng<- seq(0, 10, by=0.001)
bur<-data.frame(leng)
bur$OR<-bur$leng^0.57
bur$OR2<-1/bur$OR


ggplot(NULL, aes(x, y )) + 
  geom_smooth(data= bur, aes(x = leng, y = OR)) +  
  labs(title = "Ratio of odds of response on ratio in questionnaire lenght", x = "Ratio in lenght long to short questionnaire", y = "Ratio in odds of responding to shorter over longer questionnaire")

ggplot(NULL, aes(x, y )) + 
  geom_smooth(data= bur, aes(x = leng, y = OR2)) +  
  labs(title = "Ratio of odds of response on ratio in questionnaire lenght", x = "Ratio in lenght long to short questionnaire", y = "Ratio in odds of responding to long over short questionnaire")


##############################################################################################################
##############################################################################################################
######################################### simulation ############################################################
##############################################################################################################
##############################################################################################################


#######################################
## parameters 
#####################################

setwd("C:/Users/nf18217/OneDrive - University of Bristol/Documents/Projects/active/phil ME")
df<-read.csv("normal voc.csv")


# measurement validity
########################
## classical merriment error (i.e. reductions in validity/increases in noise or missclassification) is assumed to add a random normal variable with a 
## mean of 0 and a SD which determines the amount of noise. 
## this section allows one to model the effect of using only approximately valid measures, which is more realistic 
## than assuming that the cheaper instrument is perfectly valid.
s<-3648434384 #arbitrary number used as SD in population for validity calculation 
#e<-0.34 #for e of 0.34 approximately 95
#e<-0.48 # scaling factor between SD of population and SD of measurement error/invalidity term. for e of 0.48 this is approximately 90%
#e<-0.62 #e of 0.62 is approximately 85
#e<-0.75 # for e of 0.75 this is approximately 80%
#e<-0.88 # for e of 0.88 approximately 75
e<-1.01 # for e of 1.01 this is approximately 70%
p1<-rnorm(1000000,0,s)
p2<-p1+rnorm(1000000,0,s*e)
cor.test(p1,p2) # approximate concurrent validity of two items. 
e<-e+1 # e of 0.5 requires a 50% increase rather than dividing by two, therefore add 1 so that e*SD will reach the correct amount 

# absolute mean difference
########################
d <-0.1


# power and p-value
#####################
# beta = 0.9, alpha = 0.05; F = 10.51
# beta = 0.8, alpha = 0.05; F = 7.85
f<- 10.51

# costs
########
ce<-5 #cost of questionnaire
cg<-50 # cost of gold standard 


#####################################################################################################
#sample size and cost calculations for number of items ***assuming SD is equal in two groups***
# ***ignoring impact of questionnaire length on non-response****
#####################################################################################################

#notes
# to calculate the above I assume that the measured variance = variance in population + VOM + variance due to noise from invalidity (i.e. (SD*e)^2

#no error
df$n100<-2*f*(df$sd^2+df$vom100)/(d^2)
df$c100<-df$n100*cg

#15 items
df$n15<-2*f*(df$sd^2+df$vom15+(df$sd*e)^2)/(d^2)
df$c15<-df$n15*ce
df$d_c15<-df$c100/df$c15

#10 items
df$n10<-2*f*(df$sd^2+df$vom10+(df$sd*e)^2)/(d^2)
df$c10<-df$n10*ce
df$d_c10<-df$c100/df$c10

#8 items
df$n8<-2*f*(df$sd^2+df$vom8+(df$sd*e)^2)/(d^2)
df$c8<-df$n8*ce
df$d_c8<-df$c100/df$c8

#5 items
df$n5<-2*f*(df$sd^2+df$vom5+(df$sd*e)^2)/(d^2)
df$c5<-df$n5*ce
df$d_c5<-df$c100/df$c5

# 3 items
df$n3<-2*f*(df$sd^2+df$vom3+(df$sd*e)^2)/(d^2)
df$c3<-df$n3*ce
df$d_c3<-df$c100/df$c3

# 2 items
df$n2<-2*f*(df$sd^2+df$vom2+(df$sd*e)^2)/(d^2)
df$c2<-df$n2*ce
df$d_c2<-df$c100/df$c2


########################################################################
# drawing graphs
######################################################################


  ggplot(NULL, aes(x, y )) + 
  geom_point(data= df, aes(x = sd, y = d_c15, colour="15")) +  
  geom_point(data= df, aes(x = sd, y = d_c10, colour="10")) +
    geom_point(data= df, aes(x = sd, y = d_c8, colour="8")) +
    geom_point(data= df, aes(x = sd, y = d_c5, colour="5")) +
    geom_point(data= df, aes(x = sd, y = d_c3, colour="3")) +
  geom_point(data= df, aes(x = sd, y = d_c2, colour="2")) +
    labs(title = "Ratio of costs vs SD excluding the impact of non-response", x = "Standard deviation of population", y = "Ratio of cost of recruitment for the expensive measure/cheap measure", colour = "number of items", subtitle = "mean difference = 0.1, concurrent validity = 70%, power = 90%, alpha = 5%")
  
  
  


#####################################################################################################
#sample size and cost calculations for number of items ***assuming SD is equal in two groups***
# ***ignoring impact of questionnaire length on non-response****
#####################################################################################################

#notes
# to calculate the above I assume that the measured variance = variance in population + VOM + variance due to noise from invalidity (i.e. (SD*e)^2

#15 items
df$n15b<-2*f*(df$sd^2+df$vom15+(df$sd*e)^2)/(d^2)/(exp(0.57*log(100/15)))
df$c15b<-df$n15b*ce
df$d_c15b<-df$c100/df$c15b

#10 items
df$n10b<-2*f*(df$sd^2+df$vom10+(df$sd*e)^2)/(d^2)/(exp(0.57*log(100/10)))
df$c10b<-df$n10b*ce
df$d_c10b<-df$c100/df$c10b

#8 items
df$n8b<-2*f*(df$sd^2+df$vom8+(df$sd*e)^2)/(d^2)/(exp(0.57*log(100/8)))
df$c8b<-df$n8b*ce
df$d_c8b<-df$c100/df$c8b

#5 items
df$n5b<-2*f*(df$sd^2+df$vom5+(df$sd*e)^2)/(d^2)/(exp(0.57*log(100/5)))
df$c5b<-df$n5b*ce
df$d_c5b<-df$c100/df$c5b

# 3 items
df$n3b<-2*f*(df$sd^2+df$vom3+(df$sd*e)^2)/(d^2)/(exp(0.57*log(100/3)))
df$c3b<-df$n3b*ce
df$d_c3b<-df$c100/df$c3b

# 2 items
df$n2b<-2*f*(df$sd^2+df$vom2+(df$sd*e)^2)/(d^2)/(exp(0.57*log(100/2)))
df$c2b<-df$n2b*ce
df$d_c2b<-df$c100/df$c2b


########################################################################
# drawing graphs
######################################################################


ggplot(NULL, aes(x, y )) + 
  geom_point(data= df, aes(x = sd, y = d_c15b, colour="15")) +  
  geom_point(data= df, aes(x = sd, y = d_c10b, colour="10")) +
  geom_point(data= df, aes(x = sd, y = d_c8b, colour="8")) +
  geom_point(data= df, aes(x = sd, y = d_c5b, colour="5")) +
  geom_point(data= df, aes(x = sd, y = d_c3b, colour="3")) +
  geom_point(data= df, aes(x = sd, y = d_c2b, colour="2")) +
  labs(title = "Ratio of costs vs SD including the impact of non-response", x = "Standard deviation of population", y = "Ratio of cost of recruitment for the expensive measure/cheap measure", colour = "number of items", subtitle = "mean difference = 0.1, concurrent validity = 70%, power = 90%, alpha = 5%")


  
##############################################################################################################
##############################################################################################################
######################################### END ############################################################
##############################################################################################################
##############################################################################################################
