## Supplementary Table 4 for "Silence is golden, by my measures still see: why cheap-but-noisy outcome measures can be more cost effective than gold standards"

The formula for a sample size calculation where F is the function of the power and type one error rate, n is the required sample size in each of the two samples, σ*_i_* is the standard deviation of the outcome in the respective populations, and d is the hypothesised difference in the population outcome means:

n> F(σ_1_^2^+ σ_2_^2^)/d^2^

If we assume that the standard deviation in both populations is equal then:

n> F(σ^2^+ σ^2^)/d^2^ = 2F σ^2^/d^2^

σ is the combination of the variation of the outcome in the population [σ*_i_*], the noise (i.e. classical measurement error) in the outcome measure σ_n_, and the variance from categorising (σ_c_^2^). Therefore:

n> 2F(σ_c_^2^+ σ*_i_*^2^+ σ_n_^2^)/d^2^

If we assume that the gold standard outcome measure has no σ_c_^2^ and is measured with a perfectly valid instrument:

n_gold_ >2F σ*_i_*^2^ /d^2^

Let C_gold_ be the cost of administering the gold standard measure to one participant. The cost of the gold standard measure is:

C_gold_ *n_gold_ > C_gold_ 2F σ*_i_*^2^/d^2^

Let C_cheap_ be the cost of administering the cheaper measure. The cost of the cheaper measure is:

n_cheap_ * C_cheap_ > C_cheap_ 2F(σ_c_^2^ + σ*_i_*^2^+ σ_n_^2^)/ d^2^

We want to use the cheaper measure only when it is more cost effective. Operationalise cost effectiveness as a lower cost for the same power. Given that n is the sample size required for the desired power and type one error rate, the cheaper measure will be more cost effective when:

C_gold_ *n_gold_ > n_cheap_ * C_cheap_

Which is equivalent to:

C_gold_ 2F σ*_i_*^2^/d^2^  > C_cheap_ 2F(σ_c_^2^ + σ*_i_*^2^+ σ_n_^2^)/ d^2^

Which simplifies to:

C_gold_ / C_cheap_ > (σ_c_^2^ + σ*_i_*^2^+ σ_n_^2^)/σ*_i_*^2^

Therefore, as long as the ratio of the costs is greater than the ratio of the variances of the cheaper measure over the variance of the outcome in the population, the cheaper measure will be more cost effective.

Supplementary Table 4: Analytic conditions for when a simple measure is more cost effective
