## Supplementary Table 1 for "Silence is golden, by my measures still see: why cheap-but-noisy outcome measures can be more cost effective than gold standards"

| level | 1 | 2 | 3 | 4 | 5 | 6 | 7 | 8 | 9 | 10 | 11 | 12 | 13 | 14 | 15 |
| --- | --- | --- | --- | --- | --- | --- | --- | --- | --- | --- | --- | --- | --- | --- | --- |
| 1 item | | | | | | | | | | | | | | | |
| Range | 0-99 |  |  |  |  |  |  |  |  |  |  |  |  |  |  |
| Value | 49.5 |  |  |  |  |  |  |  |  |  |  |  |  |  |  |
| 2 items | | | | | | | | | | | | | | | |
| Range | 0-49 | 50-99 |  |  |  |  |  |  |  |  |  |  |  |  |  |
| Value | 24.75 | 74.5 |  |  |  |  |  |  |  |  |  |  |  |  |  |
| 3 items | | | | | | | | | | | | | | | |
| Range | 0-33 | 34-65 | 65-99 |  |  |  |  |  |  |  |  |  |  |  |  |
| Value | 16.5 | 49.5 | 82.5 |  |  |  |  |  |  |  |  |  |  |  |  |
| 5 items | | | | | | | | | | | | | | | |
| Range | 0-19 | 20-39 | 40-59 | 60-79 | 80-99 |  |  |  |  |  |  |  |  |  |  |
| Value | 9.5 | 29.5 | 49.5 | 69.5 | 89.5 |  |  |  |  |  |  |  |  |  |  |
| 8 items | | | | | | | | | | | | | | | |
| Range | 0-12 | 13-24 | 25-37 | 38-49 | 50-62 | 63-74 | 75-87 | 88-99 |  |  |  |  |  |  |  |
| Value | 6 | 18.5 | 31 | 43.5 | 56 | 68.5 | 81 | 93.5 |  |  |  |  |  |  |  |
| 10 items | | | | | | | | | | | | | | | |
| Range | 0-9 | 10-19 | 20-29 | 30-39 | 40-49 | 50-49 | 60-69 | 70-79 | 80-89 | 90-99 |  |  |  |  |  |
| Value | 4.5 | 14.5 | 24.5 | 34.5 | 44.5 | 54.5 | 64.5 | 74.5 | 84.5 | 94.5 |  |  |  |  |  |
| 15 items | | | | | | | | | | | | | | | |
| Range | 0-5 | 6-12 | 13-19 | 20-25 | 26-32 | 33-39 | 40-45 | 46-52 | 53-59 | 60-65 | 66-72 | 73-79 | 80-85 | 86-92 | 93-99 |
| Value | 2.5 | 9 | 16 | 22.5 | 34 | 36 | 42.5 | 49 | 56 | 62.5 | 69 | 76 | 82.5 | 89 | 96 |

Supplementary Table 1: Ranges and values of categories for questionnaire of different length.
